# Integrated Genomic Surveillance reveals extensive onward transmission of travel-imported SARS-CoV-2 infections in the community

**DOI:** 10.1101/2021.10.18.21264530

**Authors:** Torsten Houwaart, Samir Belhaj, Emran Tawalbeh, Dirk Nagels, Patrick Finzer, Yara Fröhlich, Assia Benmoumene, Dounia Asskali, Hussein Haidar, Janina von Dahlen, Jessica Nicolai, Lisa Stiller, Jacqueline Blum, Christian Lange, Carla Adelmann, Britta Schroer, Ute Osmers, Christiane Grice, Phillipp P. Kirfel, Hassan Jomaa, Daniel Strelow, Lisanna Hülse, Moritz Pigulla, Pascal Kreuzer, Alona Tyshaieva, Jonas Weber, Tobias Wienemann, Malte Kohns Vasconcelos, Katrin Hoffmann, Nadine Lübke, Sandra Hauka, Marcel Andree, Claus Jürgen Scholz, Nathalie Jazmati, Klaus Göbels, Rainer Zotz, Klaus Pfeffer, Jörg Timm, Lutz Ehlkes, Andreas Walker, Alexander T. Dilthey, German COVID-19 OMICS Initiative (DeCOI)

## Abstract

Integration of genomic surveillance with contact tracing provides a powerful tool for the reconstruction of person-to-person pathogen transmission chains. We report two large clusters of SARS-CoV-2 cases (“Delta” clade, 110 cases combined) detected in July 2021 by Integrated Genomic Surveillance in Düsseldorf. Structured interviews and deep contact tracing demonstrated an association to a single SARS-CoV-2 infected return traveller (Cluster 1) and to return travel from Catalonia and other European countries (Cluster 2), highlighting the importance of containing travel-imported SARS-CoV-2 infections.

## Main text

International travel has been shown to play an important role in the spread of SARS-CoV-2 ^1,2^. Data on the extent and structure of person-to-person SARS-CoV-2 transmission in the population following international travel introduction events, however, are scarce. We report on two large return travel-associated SARS-CoV-2 infection clusters (110 cases combined) in the populations of the German city of Düsseldorf, an international air travel and economic hub of ca 600,000 inhabitants, and a nearby smaller city, Solingen). Investigation of the underlying viral transmission chains with the Düsseldorf system of Integrated Genomic Surveillance ^3^ demonstrated that the confluence of small numbers of undetected cases in return travellers, non-adherence to infection containment measures in nightlife contexts, and subsequent community transmission can result in substantial numbers of SARS-CoV-2 infections.

### Integrated Genomic Surveillance in Düsseldorf

The system of Integrated Genomic Surveillance (IGS) in the city of Düsseldorf has been described elsewhere ^3^. Briefly, the system involves rapid sequencing of a large proportion of local SARS-CoV-2 cases at the Centre for Medical Microbiology, Hospital Hygiene, and Virology of Heinrich Heine University Düsseldorf and an integration of genetic data with intensified backward contact tracing and structured case interviews at Düsseldorf Health Authority for the improved characterization of population transmission chains.

Between 15 June and 01 August 2021, the system generated 518 high-quality SARS-CoV-2 genome sequences (< 5000 undefined nucleotides; Supplementary Table 1); during the same period, 976 SARS-CoV-2 cases were registered in Düsseldorf. The system’s routine cluster analysis algorithms, based on the identification of groups of pairwise-identical viral isolates (“cliques”), suggested that two large novel clusters of closely related viral isolates, belonging to the “Delta” clade ^4^, had appeared in July 2021. Phylogenetic analysis (Supplementary Figure 1; Supplementary Note) was used to confirm and refine the definition of these clusters. To investigate potential transmission beyond Düsseldorf, available sequences from the nearby city of Solingen, where a trial run of the IGS system took place in July and August 2021, were also integrated into the sample. Cluster 1 consisted of 60 viral sequences with an average pairwise genetic distance (defined in Supplementary Note) of 0.91 (59 from Düsseldorf, 1 from Solingen), sampled from 05 July onwards (Supplementary Table 2); Cluster 2, of 42 viral sequences with an average pairwise genetic distance of 1.89 (36 from Düsseldorf, 6 from Solingen), sampled from 30 June onwards (Supplementary Table 3).

To investigate the origin of these clusters, contact tracing data routinely collected at Düsseldorf and Solingen Health Authorities were integrated with structured interviews of the Düsseldorf cases (“deep backward contact tracing”), covering (i) occupation and place of work; (ii) utilization of public transport; (iii) social, household and family contacts; (iv) utilization of medical services; (v) supermarket and retailer visits; (vi) gastronomy and nightlife; (vii) travel history.

### Cluster 1 originated from a single infected case followed by nightlife spreading events

The emergence of Cluster 1 in Düsseldorf could be traced back to an individual SARS-CoV-2-infected traveller returning to Düsseldorf from the island of Mallorca (“IP”) on day 0; the identified links between samples are visualized in Figure 1. Transmission of the imported viral strain (“Delta” clade) in Düsseldorf was initiated when IP and eight first-order contacts (KP1 – KP8) visited two bars (“Bar A”, “Bar B”) in a popular nightlife of Düsseldorf, an area with narrow streets and more than 200 bars, on day 2. Additional transmissions took place in a complex pattern of additional visits of the first-order contacts of IP to Bar A on day 4 and day 5 (Figure 1B Inset); during a likely encounter between the first-order contacts of IP and KP9 and KP10, who were on a pub crawl in the area around Bar A on day 5; and from the second-order contacts of IP into the local population via private meetings, family and household contacts (Figure 1B). Investigations by Düsseldorf Health Authority showed limited adherence to SARS-CoV-2 infection prevention rules and measures in Bar A and B, including dancing without masks and insufficient tracking of customer contact details. Including IP and 5 individuals who were in the company of IP and KP1 during their visits to Bar A, who later tested positive for SARS-CoV-2, and the viral sequences of whom were not available, 28 SARS-CoV-2 cases in Düsseldorf could be directly linked to IP (Figure 1A). Contact tracing and structured interviews also uncovered links between an additional 15 cases without direct links to IP; these likely represented ongoing community transmission of the introduced viral strain (Figure 1B). The assignment of IP as the index case was supported by date of symptom onset (Supplementary Table 2) and by the fact that a GISAID Audacity Instant search for the viral sequence of IP (which was determined by a commercial diagnostic laboratory and obtained via GISAID ^5^; accession EPI_ISL_3044996) identified a closely related sequence (accession EPI_ISL_2710175) from the Balearic Islands with genetic distance 1, collected ca. two weeks prior to IP’s return from Mallorca and lacking a defining mutation of the Cluster 1 sequences (Supplementary Note). Including IP and the 5 epidemiologically linked first- and second-order contacts of IP (see above), Cluster 1 comprised 67 Düsseldorf cases, or 8% of new SARS-CoV-2 cases registered in Düsseldorf in July (Figure 2). Of note, two cases belonging to Cluster 2, Z4187 and Z4145, also visited Bar A on day 4; despite this potential epidemiological link, the genetic data clearly showed that these belonged to a different infection cluster. For S88, the only Solingen sample in Cluster 1, no links to the Düsseldorf cases or other putative infection sources were identified.

**Figure 1:**
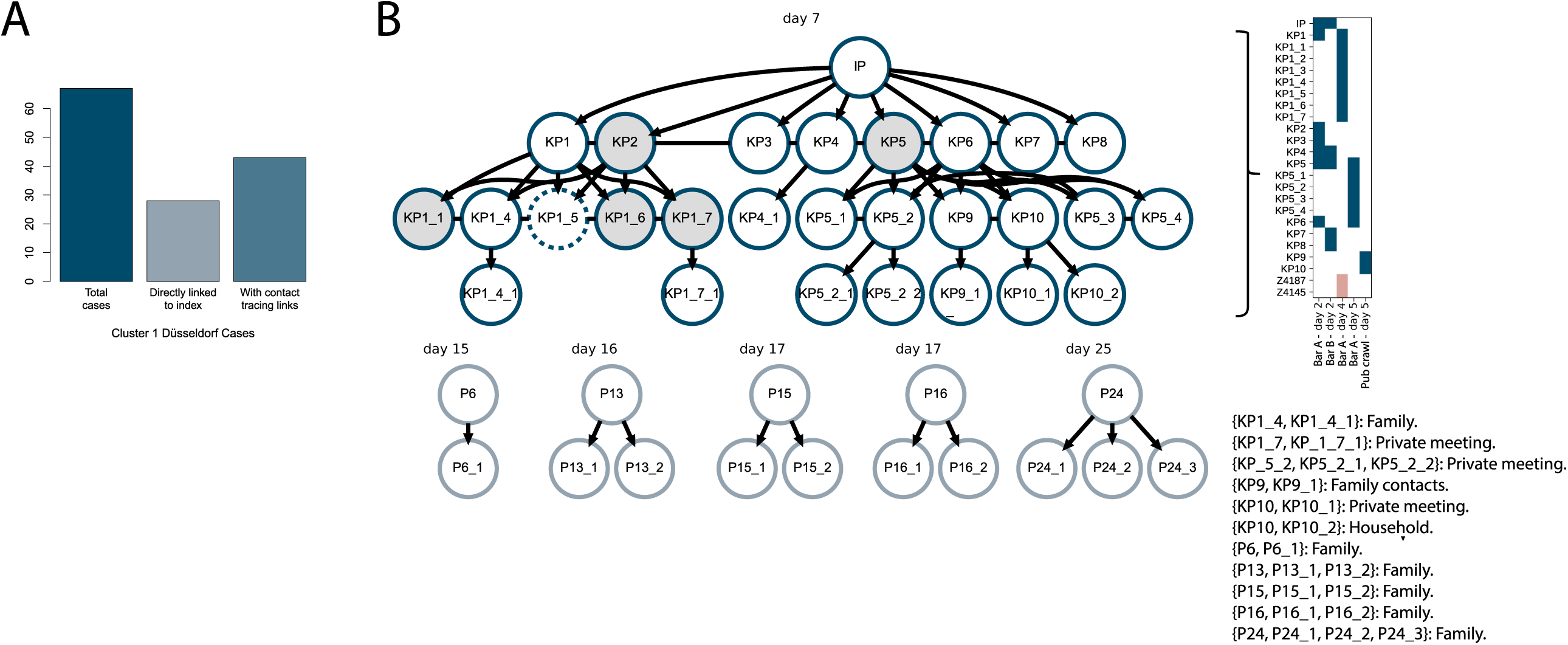
Cluster 1 contact tracing results. **A**. Bar plot showing the total number of cases in Cluster 1 (after inclusion of cases directly linked to IP and KP1, see text), how many of these were directly linked to the index case, and for how many of these routine contact tracing and structured interviews uncovered links to other Cluster 1 samples. **B**. Visualization of the reconstructed epidemiological structure of Cluster 1. Each node in the transmission chain graph represents one case. Nodes shaded in gray represent epidemiologically linked, but non-sequenced cases that were in the direct company of IP or KP1 during their visits to Bar A. Test dates of the assumed index cases of the connected components are shown above the nodes representing the corresponding cases. The inset to the right of the transmission chain graph shows the complex patterns of visits of IP and their first- and second-order contacts to two bars in the nightlife district of Düsseldorf. The two cases shown with red boxes also visited Bar A, but their sequenced viral isolates cluster with Cluster 2. Samples KP9 and KP10 participated in a pub crawl in the same area around Bar A. The text inset details the precise nature of the identified case relationships. KP1_5 exhibited an increased genetic distance to the other samples in the cluster and is therefore shown with a dashed border; see Supplementary Note for a discussion. Transmission chain graphs were plotted with Graphviz ^6^.

**Figure 2:**
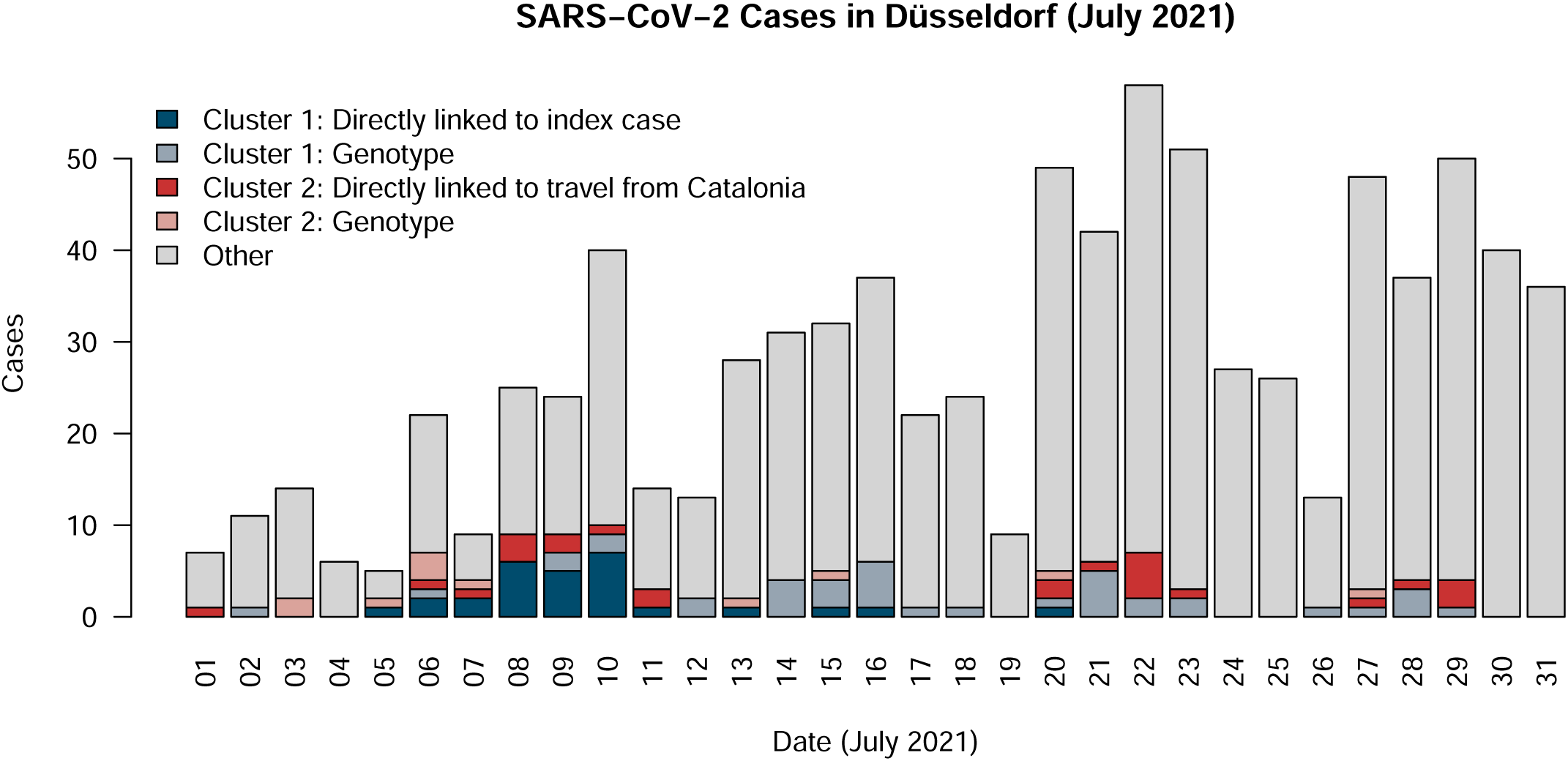
SARS-CoV-2 cases in Düsseldorf in July 2021. The figure shows daily new registered SARS-CoV-2 cases in Düsseldorf in July 2021, and how many of these were associated with Cluster 1 and Cluster 2. For Cluster 1, “directly linked to index case” refers to uninterrupted, contact tracing-supported potential transmission chains between the linked cases and the index patient; “genotype” refers to samples that were identified as belonging to Cluster 1 in the phylogenetic tree analysis (see text). For Cluster 2, “directly linked to travel from Catalonia” refers to cases that either have recently returned from Catalonia and to cases that are directly linked to the Catalonia returnees in a manner supported by contact tracing; the definition of “genotype” is identical to that for Cluster 1.

### Cluster 2 represents multiple independent importation events linked to return travel

For Cluster 2 no clear index could be identified. While intensified contact tracing and structured case interviews enabled delineating relationships for 25 cases (e.g. household contacts), the overall size of the identified transmission chains was limited in size compared to Cluster 1 (Figure 3A). Examination of the travel history of the cases, however, showed that almost a quarter of Cluster 2 cases could be linked to return travel from Catalonia (7 returnees from Catalonia and 3 associated downstream infections in Düsseldorf); an additional 5 cases had been to France prior to testing positive for SARS-CoV-2 (Figure 3B, Supplementary Table 3). Analysis of symptom onset (Supplementary Table 3) in relation to return travel dates showed that the majority of return travellers in Cluster 2 were likely infected during their stay abroad; for some cases, exposure in Düsseldorf could be ruled out with certainty (e.g. Z4106, with symptom onset on the same day as the return flight). The viral sequences of Z4077 and Z4076, who were among the earliest Cluster 2 cases in Düsseldorf and who were likely infected during a joint trip to Barcelona (symptom onset 1 and 3 days after return to Düsseldorf, respectively), exhibited a genetic distance of 3, indicating two independent infection events. Cluster 2 thus likely reflected multiple independent introduction events of a viral strain also circulating in Catalonia and other European countries, followed by diffuse community transmission in Düsseldorf. Of note, an additional direct contact of Z4077 and Z4076 in Barcelona also tested positive for SARS-CoV-2 after return to Düsseldorf and reported many contacts at work and in social gatherings in Düsseldorf and other cities, possibly contributing to community transmission; the viral sequence of this case could be obtained through a commercial diagnostic lab in Cologne and was found to also cluster with Cluster 2 (data not shown). Consistent with diffuse community transmission in Düsseldorf and surrounding cities, for 5 additional cases from Düsseldorf, and 1 case from Solingen at the beginning of a six-person transmission chain, case interviews suggested a possible exposure to the virus in the nightlife district of Düsseldorf (Figures 3A and 3B). Furthermore, consistent with circulation of the Cluster 2 viral strain in other European countries, sequence-identical samples from multiple European countries were identified via a GISAID Audacity Instant search for the sequence of Z4077, with sample collection dates of the identified GISAID beginning on 29 June, i.e. nearly identical to the sample collection dates of the first Cluster 2 cases in Düsseldorf. Over the course of July, Cluster 2 accounted for 4% of total SARS-CoV-2 infections in Düsseldorf.

**Figure 3:**
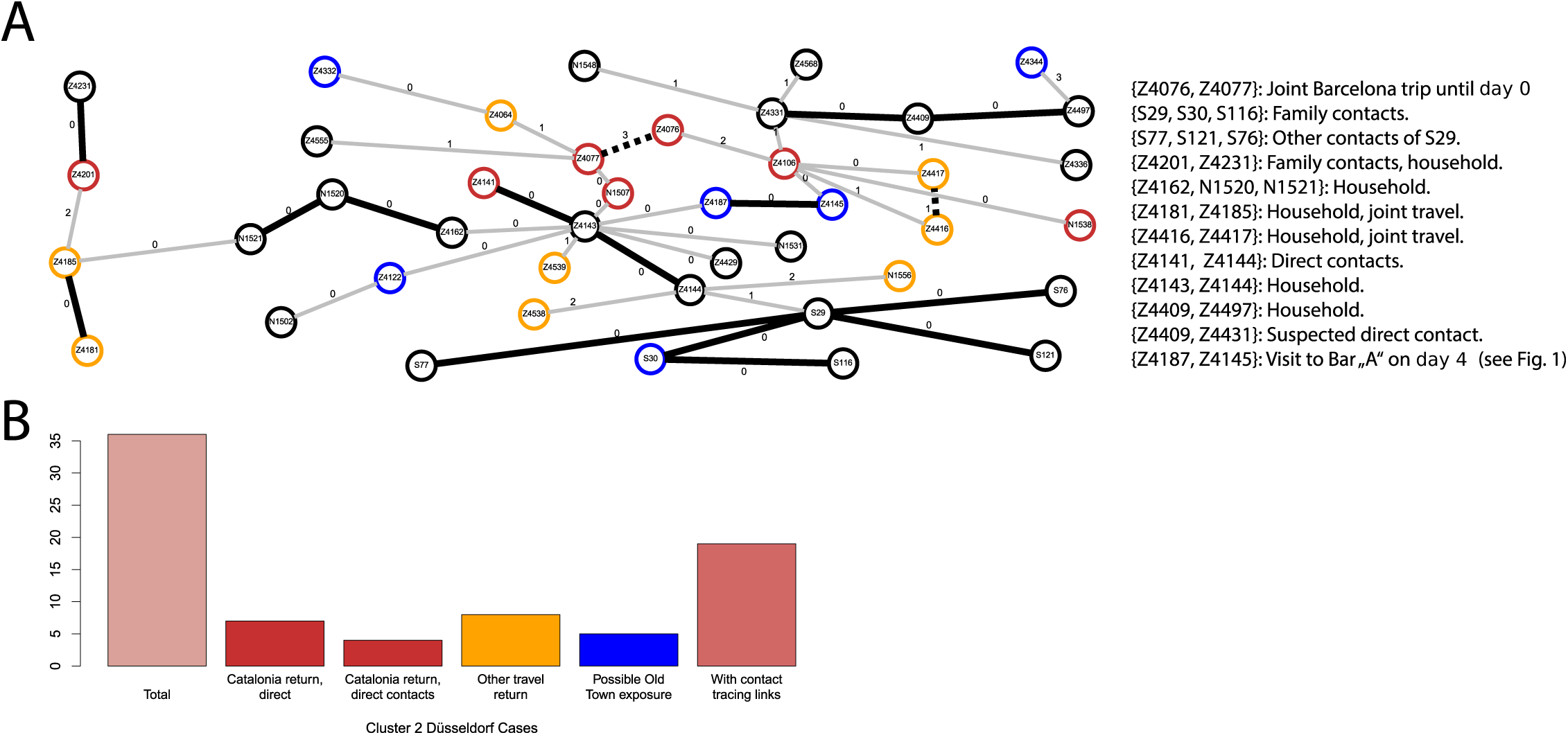
Cluster 2 genetic structure and contact tracing results. **A**. Integrated visualization of genetic distances and contact tracing results for Cluster 2. The visualization ^6^ is based on a Minimum Spanning Tree (MST) of the genetic distances between samples, selecting the tree that contains the highest number of edges also present in the contact tracing data from the set of optimal (minimum cumulative edge weight) trees. Each node represents one case; each edge is labelled with the genetic distance between the two samples it connects. Black edges represent links also present in the contact tracing data; grey edges represent links not present in the contact tracing data. Dashed black edges represent links present in the contact tracing data, but not in the MST. Node colours (see figure part B) indicate recent travel history and possible exposure in the nightlife distritct of Düsseldorf. **B**. Bar plot showing the total number of cases in Cluster 2, for how many of these a travel history background or a direct link to a recently returned case from Catalonia could be established, and for how many of these routine contact tracing and structured interviews uncovered links to other Cluster 2 samples.

## Discussion and Conclusion

The investigated clusters demonstrate the potential of Integrated Genomic Surveillance to characterize person-to-person SARS-CoV-2 transmission in the population and provide clear evidence for the importance of the containment of travel-imported SARS-CoV-2 infections.

A large proportion of cases in Cluster 1 could be directly associated with a single undetected SARS-CoV-2 infection in a return traveller; subsequent spread in the community was mediated by non-adherence to infection control and prevention measures in nightlife venues, and tracing and containment efforts by Düsseldorf Public Health Authorities were hampered by insufficient tracking of customer contact details. Cluster 2, by contrast, was driven by multiple independent introductions of a viral strain also circulating in Catalonia and other parts of Europe, and found in another city, Solingen, of the wider metropolitan area around Düsseldorf.

Resolving the two clusters would not have been possible without IGS; in many instances, links between cases were only uncovered by the structured case interviews carried out after genetic links had been identified. In addition, due to epidemiological links between the two clusters (samples Z4187 and Z4145), Cluster 1 and 2 would have been considered as connected without the additional information gathered through Integrated Genomic Surveillance.

Nightclubs are generally prone to accumulate large amounts of potentially infectious aerosols and visitors are likely to neglect individual infection control measures; limiting entry to such venues to people immunized through vaccination or previous infection or those with a current PCR test could drastically limit the risk of transmission and would likely have contributed to a reduction of cases in Cluster 1.

Spain, including the Balearic Islands, was declared a region of high Covid-19 risk on July 27 ^7^, i.e. almost 4 weeks after the first transmission in the reported clusters occurred. No mandatory quarantine measures were in place for travellers returning from this region at that time. Strict PCR testing of travellers both at the time of return and after a mandatory quarantine could have contributed to a significant reduction of cases in Cluster 1 and 2 and reduced overall case load in Düsseldorf in July 2021 (Figure 2).

## Data Availability

FASTA files and the phylogenetic tree of all Düsseldorf and Solingen samples have been made available via OSF (DOI 10.17605/OSF.IO/B7TU3). All viral genome assemblies from Düsseldorf surveillance samples that meet GISAID quality criteria have also been submitted to GISAID. Accessions are listed in Supplementary Table 1.

https://osf.io/b7tu3/

https://www.gisaid.org/

## Acknowledgements and Data Availability

This work was supported by the Ministry for Work, Health and Social Affairs of the State of North Rhine-Westphalia; the Jürgen Manchot Foundation; the German Federal Ministry of Education and Research (Bundesministerium für Bildung und Forschung; award numbers 031L0184B and 01KX2021); the German Research Foundation (award 428994620).

We gratefully acknowledge the Authors, the Originating and Submitting Laboratories for their sequence and metadata shared through GISAID. All submitters of data may be contacted directly via <u>GISAID</u>. The Acknowledgments Table for GISAID is part of the Supplement (Supplementary Table S4). Computational support and infrastructure were provided by the “Centre for Information and Media Technology” (ZIM) at the University of Düsseldorf (Germany). Sequencing and the design of sequencing workflows were supported by the Biologisch-Medizinisches Forschungszentrum der Heinrich Heine University Düsseldorf (BMFZ).

## Supplementary Table Legends

**Supplementary Table S1: Sample and sequencing data summary**. The table summarizes sequencing data statistics, assembly quality, variant calls, and accession numbers for the samples of this study.

**Supplementary Table S2: Cluster 1 samples**. For each sample in Cluster 1, the table shows the dates of the first positive test and of symptom onset (if any), the mapping between FASTA and contact tracing IDs, whether a case is directly linked to the index case IP and whether a case belongs to the city of Düsseldorf (columns “DirectlyLinked” and “Case from Dusseldorf”), the genetic distance to case IP (column “distanceToIndex”), and details on the results of the integration of routine contact tracing and the structured case interviews.

**Supplementary Table S3: Cluster 2 samples**. For each case in Cluster 2, the table shows the dates of the first positive test and of symptom onset (if any), whether a case is directly linked to return travel from Catalonia (column “DirectlyLinked”), whether a case has a recent travel history to Catalonia (column “ReturneeFromCatalonia”), whether a case has a recent travel history to other countries (column “ReturneeFromElsewhere”), whether a case belongs to the city of Düsseldorf (column “Case from Dusseldorf”), whether a case could derive from exposure in nightlife district of Düsseldorf (column “Possible exposure in nightlife district”), the genetic distance to case Z4077, one of the earliest Cluster 2 cases in Düsseldorf (column “Genetic distance to Z4077”), and details on the results of the integration of routine contact tracing and the structured case interviews.

**Supplementary Table S4: Acknowledgements table for GISAID**.

**Supplementary Figure 1:** iTol ^8^ visualization of the phylogenetic tree used for defining Cluster 1 and Cluster 2 (Supplementary Note).

**Supplementary Note:** Full descriptions of the definitions of Cluster 1 and Cluster 2; of the genetic distance calculation; discussion of KP1_5.

## German COVID-19 OMICs Initiative (DeCOI)

Janine Altmüller, Angel Angelov, Anna C. Aschenbrenner, Robert Bals, Alexander Bartholomäus, Anke Becker, Matthias Becker, Daniela Bezdan, Michael Bitzer, Conny Blumert, Ezio Bonifacio, Peer Bork, Bunk Boyke, Helmut Blum, Nicolas Casadei, Thomas Clavel, Maria Colome-Tatche, Markus Cornberg, Inti Alberto De La Rosa Velázquez, Andreas Diefenbach, Alexander Dilthey, Nicole Fischer, Konrad Förstner, Sören Franzenburg, Julia-Stefanie Frick, Gisela Gabernet, Julien Gagneur, Tina Ganzenmueller, Marie Gauder, Janina Geißert, Alexander Goesmann, Siri Göpel, Adam Grundhoff, Hajo Grundmann, Torsten Hain, Frank Hanses, Ute Hehr, André Heimbach, Marius Hoeper, Friedemann Horn, Daniel Hübschmann, Michael Hummel, Thomas Iftner, Angelika Iftner, Thomas Illig, Stefan Janssen, Jörn Kalinowski, René Kallies, Birte Kehr, Andreas Keller, Oliver T. Keppler, Sarah Kim-Hellmuth, Christoph Klein, Michael Knop, Oliver Kohlbacher, Karl Köhrer, Jan Korbel, Peter G. Kremsner, Denise Kühnert, Ingo Kurth, Markus Landthaler, Yang Li, Kerstin U. Ludwig, Oliwia Makarewicz, Manja Marz, Alice C. McHardy, Christian Mertes, Maximilian Münchhoff, Sven Nahnsen, Markus Nöthen, Francine Ntoumi, Peter Nürnberg, Stephan Ossowski, Jörg Overmann, Silke Peter, Klaus Pfeffer, Isabell Pink, Anna R. Poetsch, Ulrike Protzer, Alfred Pühler, Nikolaus Rajewsky, Markus Ralser, Kristin Reiche, Olaf Rieß, Stephan Ripke, Ulisses Nunes da Rocha, Philip Rosenstiel, Antoine-Emmanuel Saliba, Leif Erik Sander, Birgit Sawitzki, Simone Scheithauer, Philipp Schiffer, Jonathan Schmid-Burgk, Wulf Schneider, Eva-Christina Schulte, Joachim L. Schultze, Alexander Sczyrba, Mariam L. Sharaf, Yogesh Singh, Michael Sonnabend, Oliver Stegle, Jens Stoye, Fabian Theis, Thomas Ulas, Janne Vehreschild, Thirumalaisamy P. Velavan, Jörg Vogel, Sonja Volland, Max von Kleist, Andreas Walker, Jörn Walter, Dagmar Wieczorek, Sylke Winkler, John Ziebuhr

