## Supplementary figures and images for "Integrated Genomic Surveillance reveals extensive onward transmission of travel-imported SARS-CoV-2 infections in the community"

### Supplementary Figure 1

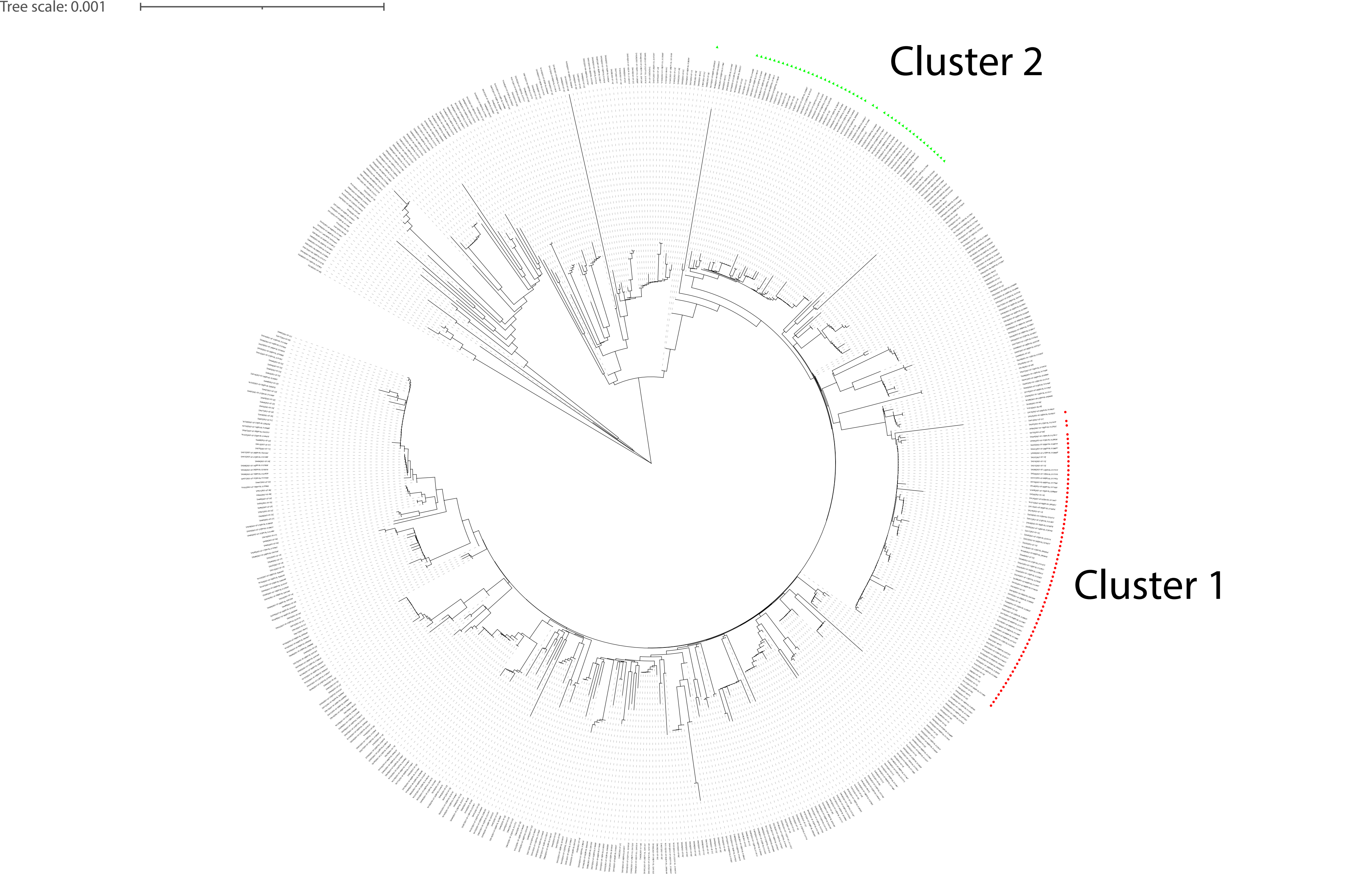
