## Supplementary Note for "Integrated Genomic Surveillance reveals extensive onward transmission of travel-imported SARS-CoV-2 infections in the community"

### Phylogenetic analysis and definition of C1 and C2

A phylogenetic tree of all sequences registered by the Integrated Genomic Surveillance (IGS) platform of Düsseldorf and sampled between 15 June 2021 and 01 August 2021 was constructed using the neighbour-joining method with the Tamura-Nei genetic distance model as implemented in Geneious version 10.2.6. The FASTA file used for the phylogenetic analysis as well as the constructed tree in NEWICK format are provided via the Open Science Foundation; see the main text for the corresponding accession numbers. An iTol [Letunic and Bork 2016] visualization of the phylogenetic tree (including the defined clusters, see below) is shown in Supplementary Figure 1.

The samples previously flagged by the IGS routine cluster analysis algorithms were located in the tree and the presence of two large infection clusters was confirmed (see Supplementary Figure 1). We carried out a mutational analysis and identified T14064C (red circles) as being consistently present in C1 isolates, and C18744T (green right pointing triangle) as being consistently present in C2 isolates.

Based on this analysis, we chose I361 as the root node for C1, and I584 as the root node for C2, and we defined C1 and C2 as the set of all children of these nodes, with the following exceptions:

- From C1, we removed:
  - all Düsseldorf samples that were not part of our genomic surveillance program (e.g. samples taken from Düsseldorf University Hospital inpatients, sample ID V*; none of these predated the surveillance samples).
  - Z4525, based on its identification as an outlier in a visual inspection of the tree
  - N1515, based on its identification as an outlier in a visual inspection of the tree
  - Z4412, because it had more than 5000 Ns (13842)
  - Z4153, because it had more than 5000 Ns (6507)
  - Z4398, N1509, Z4578, Z4342, because they belonged to repeat tests of cases already part of the cluster
    - Z4398 belongs to the same case as Z4262.
    - N1509 belongs to the same case as Z4206.
    - Z4578 and Z4342 belong to the same case as Z4234.
- From C2, we removed:
  - all Düsseldorf samples that were not part of our genomic surveillance program (e.g. samples taken from Düsseldorf University Hospital inpatients, sample ID V*; none of these predated the surveillance samples).
  - Z4486, because it had more than 5000 Ns (13842)
  - Z4545, Z4619, Z4424, Z4364, because they belonged to repeat tests of cases already part of the cluster
    - Z4545 belongs to the same case as Z4201.
    - Z4619 belongs to the same case as Z4231.
    - Z4424 and Z4364 belong to the same case as Z4145

The final sequence ID lists for the two clusters were:

- C1:
  - 'Z4116|2021-07-05|EPI ISL 3116653'
  - 'Z4124|2021-07-07|EPI ISL 3117447'
  - 'Z4265|2021-07-16|EPI ISL 3127825'
  - 'Z4218|2021-07-13|'
  - 'Z4550|2021-07-27|EPI ISL 3210115'
  - 'Z4528|2021-07-26|EPI ISL 3210078'
  - 'Z4295|2021-07-16|EPI ISL 3128146'
  - 'Z4217|2021-07-13|'
  - 'Z4549|2021-07-27|EPI ISL 3210114'
  - 'Z4411|2021-07-21|EPI ISL 3131487'
  - 'Z4527|2021-07-26|EPI ISL 3210077'
  - 'Z4151|2021-07-09|EPI ISL 3117470'
  - 'Z4150|2021-07-09|EPI ISL 3117469'
  - 'Z4140|2021-07-06|EPI ISL 3117459'
  - 'Z4117|2021-07-06|EPI ISL 3116654'
  - 'Z4365|2021-07-21|EPI ISL 3128626'
  - 'Z4352|2021-07-20|EPI ISL 3128614'
  - 'Z4341|2021-07-20|EPI ISL 3128601'
  - 'Z4340|2021-07-21|EPI ISL 3128600'
  - 'Z4237|2021-07-14|'
  - 'Z4216|2021-07-13|'
  - 'Z4215|2021-07-13|'
  - 'Z4206|2021-07-12|EPI ISL 3117513'
  - 'Z4203|2021-07-12|EPI ISL 3117510'
  - 'Z4226|2021-07-14|'
  - 'Z4254|2021-07-15|EPI ISL 3127817'
  - 'Z4107|2021-07-08|EPI ISL 3116639'
  - 'Z4188|2021-07-09|EPI ISL 3117497'
  - 'Z4234|2021-07-14|'
  - 'Z4232|2021-07-14|'
  - 'Z4361|2021-07-22|EPI ISL 3128623'
  - 'Z4442|2021-07-22|'
  - 'Z4360|2021-07-22|EPI ISL 3128622'
  - 'Z4397|2021-07-21|EPI ISL 3131470'
  - 'Z4174|2021-07-09|EPI ISL 3117487'
  - 'Z4152|2021-07-09|EPI ISL 3117471'
  - 'Z4621|2021-08-01|'
  - 'Z4551|2021-07-27|EPI ISL 3210117'
  - 'Z4573|2021-07-28|EPI ISL 3247243'
  - 'Z4292|2021-07-17|EPI ISL 3127845'
  - 'Z4179|2021-07-09|EPI ISL 3117491'
  - 'Z4176|2021-07-09|EPI ISL 3117489'
  - 'Z4173|2021-07-09|EPI ISL 3117486'
  - 'Z4154|2021-07-09|EPI ISL 3117472'
  - 'Z4255|2021-07-15|'
  - 'N1518|2021-07-11|EPI ISL 2993654'
  - 'Z4449|2021-07-23|'
  - 'Z4261|2021-07-15|EPI ISL 3127821'
  - 'Z4353|2021-07-20|EPI ISL 3128615'
  - 'Z4262|2021-07-15|EPI ISL 3127822'
  - 'N1501|2021-07-07|EPI ISL 2993637'
  - 'S88|2021-07-14|'
  - 'Z4274|2021-07-17|EPI ISL 3127830'
  - 'Z4339|2021-07-20|EPI ISL 3128599'
  - 'N1506|2021-07-09|EPI ISL 2993642'
  - 'N1524|2021-07-20|EPI ISL 3144471'
  - 'Z4202|2021-07-13|'
  - 'Z4260|2021-07-15|'
  - 'Z4177|2021-07-09|EPI ISL 3117490'
  - 'Z4155|2021-07-09|'
- C2:
  - 'Z4106|2021-07-05|EPI ISL 3116638'
  - 'Z4143|2021-07-06|EPI ISL 3117462'
  - 'Z4145|2021-07-08|EPI ISL 3117465'
  - 'Z4144|2021-07-06|EPI ISL 3117463'
  - 'Z4181|2021-07-09|EPI ISL 3117493'
  - 'Z4162|2021-07-09|EPI ISL 3117478'
  - 'Z4231|2021-07-14|'
  - 'Z4201|2021-07-13|EPI ISL 3117509'
  - 'Z4185|2021-07-09|EPI ISL 3117495'
  - 'Z4538|2021-07-26|EPI ISL 3210094'
  - 'Z4187|2021-07-08|EPI ISL 3117496'
  - 'N1521|2021-07-08|EPI ISL 2993657'
  - 'N1520|2021-07-08|EPI ISL 2993656'
  - 'Z4076|2021-07-02|EPI ISL 3116609'
  - 'Z4429|2021-07-21|EPI ISL 3131767'
  - 'Z4077|2021-07-02|EPI ISL 3116610'
  - 'Z4417|2021-07-21|EPI ISL 3131494'
  - 'Z4539|2021-07-26|EPI ISL 3210096'
  - 'S76|2021-07-14|'
  - 'S77|2021-07-16|'
  - 'S30|2021-07-09|'
  - 'S116|2021-07-16|'
  - 'S29|2021-07-08|'
  - 'S121|2021-07-22|'
  - 'Z4064|2021-06-30|EPI ISL 3115813'
  - 'Z4332|2021-07-19|EPI ISL 3128592'
  - 'Z4141|2021-07-06|EPI ISL 3117460'
  - 'Z4122|2021-07-06|EPI ISL 3116660'
  - 'N1531|2021-07-21|EPI ISL 3144478'
  - 'N1507|2021-07-06|EPI ISL 2993643'
  - 'N1538|2021-07-20|EPI ISL 3144485'
  - 'N1556|2021-07-28|EPI ISL 3266257'
  - 'N1502|2021-07-07|EPI ISL 2993638'
  - 'Z4555|2021-07-27|EPI ISL 3210123'
  - 'Z4416|2021-07-21|EPI ISL 3131493'
  - 'Z4344|2021-07-21|EPI ISL 3128607'
  - 'Z4568|2021-07-28|EPI ISL 3247238'
  - 'Z4331|2021-07-19|EPI ISL 3128591'
  - 'Z4409|2021-07-21|EPI ISL 3131486'
  - 'Z4497|2021-07-23|'
  - 'N1548|2021-07-27|EPI ISL 3266249'
  - 'Z4336|2021-07-19|EPI ISL 3128596'

### Discussion of KP1_5

With a genetic distance of 15 to the sequence of case IP of Cluster 1, the genome sequence of KP1_5 (N1515), despite clustering with Cluster 1 isolates, was removed as an outlier during the phylogenetic analysis. The sample was externally sequenced by one of the participating commercial diagnostic labs and the detected variants include an unusual pattern of two deletions of 6 nucleotides each, possibly indicating reduced quality of the assembled genomic sequence. The case was re-integrated into the analysis as a direct contact of KP_1 and KP_2. Despite its high genetic distance of 15, the viral sequence of KP1_5 carries the mutation T14064C, which is associated with Cluster 1 in the examined dataset, and which distinguishes the sequence of IP from that of the identified genome from the Balearic Islands (GISAID accession EPI_ISL_2710175). In addition, increased genetic distances are sometimes observed in the analysis of epidemiologically clearly linked outbreak isolates [Walker et al. 2020, Walker et al. 2021]. The genetic data are therefore ambiguous with respect to the question whether sample KP1_5 belongs to the transmission chain originating with IP.

### Calculation of sample genetic distances

Genetic distances between samples were calculated as defined previously, with one modification (point d) below). Briefly, a Multiple Sequence Alignment (MSA) of all sequences was built using mafft [Katoh and Standley 2013], using GISAID instructions. The distance $d(x, y)$between two samples x and y was defined as the number of differences between the MSA entries of $x$ and $y$, a) ignoring leading or trailing gap characters, b) counting matches and mismatches according to IUPAC ambiguity codes, c) counting subsequent non-matching gaps columns as a difference of 1, d) ignoring deletions aligned to ‘N’ regions in the other genome, and e) ignoring any mismatches in the MSA regions between i) the beginning of the MSA and the 20-th ACGT character of either sequence and ii) the end of the MSA and the 20 last ACGT characters of either sequence.
